# Multi-Hospital Electronic Health Record Foundation Models Without Data Sharing: A Comparison of Federated Learning and Inference-Time Ensembling

**DOI:** 10.64898/2026.04.24.26351702

**Authors:** Olivier Elemento

## Abstract

**Background:** Foundation models for electronic health records (EHRs) perform strongly on clinical prediction, but every published model has been trained within a single health system. No multi-institutional EHR foundation model currently exists, largely because privacy regulations and governance barriers block data pooling across hospitals. Two strategies could build such models without pooling: federated learning (exchanges model weights) and inference-time ensembling (exchanges only predictions at query time). Whether either is viable for autoregressive EHR foundation models, and whether individual hospitals benefit from participating, is not established.

**Methods:** We trained a generative pretrained transformer (GPT) style EHR foundation model on 100,163 Medical Information Mart for Intensive Care (MIMIC-IV) patients, partitioned into five heterogeneously distributed (non-IID) sites by Dirichlet allocation over International Classification of Diseases (ICD) chapters. We compared centralized training, federated averaging, and inference-time ensembling, and each hospital’s solo model against the ensemble including it. Models were evaluated on 15,012 held-out patients using per-condition area under the receiver operating characteristic curve (AUROC) for five acute conditions and micro-averaged area under the precision-recall curve (AUPRC) across 2,590 diagnoses.

**Results:** Centralized training achieved per-condition AUROC 0.75–0.85 and overall AUPRC 0.376. Federated averaging recovered 85% of centralized AUPRC (0.321) and 98–100% of per-condition AUROC. Inference-time ensembling, requiring no training-time exchange, recovered 77% of AUPRC (0.291) and 97–99% of per-condition AUROC. An estimated 87% of participating hospitals received a better model from the ensemble than from training alone; only hospitals with ^∼^40% of the network’s patients matched the ensemble on their own. FedProx collapsed to the marginal baseline.

**Conclusions:** Multi-institutional EHR foundation models can be built without pooling patient data. Inference-time ensembling benefits most participating hospitals and imposes the lightest governance burden; federated learning recovers more performance but requires weight sharing. These findings offer a practical path toward collaborative clinical AI.

## INTRODUCTION

Foundation models for electronic health records (EHRs) have emerged as a leading approach for clinical prediction. Recent work has produced institution-scale EHR foundation models at NYU Langone (1.29 million patients) (1), UCLA Health (2.4 million patients) (2), Columbia University (2.6 million patients) (3), and King’s College Hospital NHS Foundation Trust together with South London and Maudsley NHS Foundation Trust (approximately 811,000 patients) via the Foresight model (4), each demonstrating that a single GPT-style decoder trained on longitudinal patient records can perform zero-shot disease onset prediction, readmission risk estimation, and mortality forecasting across many downstream tasks. A recent benchmark of six EHR foundation models across 14 clinical prediction tasks found that GPT-style architectures consistently ranked highest on discrimination metrics (5).

Every published EHR foundation model to date has been trained on data from a single health system. No multi-institutional EHR foundation model currently exists. This is not primarily a technical gap but a governance gap: pooling EHR data across institutions is hampered in practice by the Health Insurance Portability and Accountability Act (HIPAA), institutional review boards (IRBs), state privacy laws, and contractual data-use restrictions, and is further constrained by hospitals’ legitimate concerns about liability, data ownership, and the difficulty of reversing consent once data has moved. Even research consortia and commercial healthcare AI partners that could in principle aggregate data across hospitals rarely do so for foundation-model pretraining. The situation is unlikely to change in the near term.

This absence has concrete consequences. A foundation model trained on one tertiary academic medical center may serve that hospital’s population well but generalize poorly to community hospitals with different patient mixes, documentation practices, or coding conventions. The FoMoH benchmark specifically documented this transportability gap, showing that foundation models trained at one institution degrade when evaluated at another, even when both sites use the standardized Observational Medical Outcomes Partnership (OMOP) common data model (5). A collaborative multi-institutional model would reduce this gap, improve rare-disease coverage, and produce better predictions for every participating hospital. But to be adopted, such a model must be trainable without transferring patient records.

Two technical strategies can in principle build multi-institutional foundation models without pooling data (**Figure 1**). In federated learning, each hospital trains a local model on its own data and exchanges model parameter updates with a central aggregator across training rounds, typically through weighted averaging (**Figure 1b**) (6,7). Federated learning avoids transferring patient data but still requires institutions to share model weights, which encode information about the training distribution and may be subject to the same governance concerns as the underlying data. In inference-time ensembling, each hospital trains its own model independently with no training-time communication; at query time, an inference broker forwards the query patient to each participating hospital’s model and averages their prediction probabilities (**Figure 1c**). Ensembling eliminates training-time parameter exchange entirely: nothing crosses institutional boundaries during model development, and only soft predictions on individual query patients cross at inference time.

**Figure 1.**
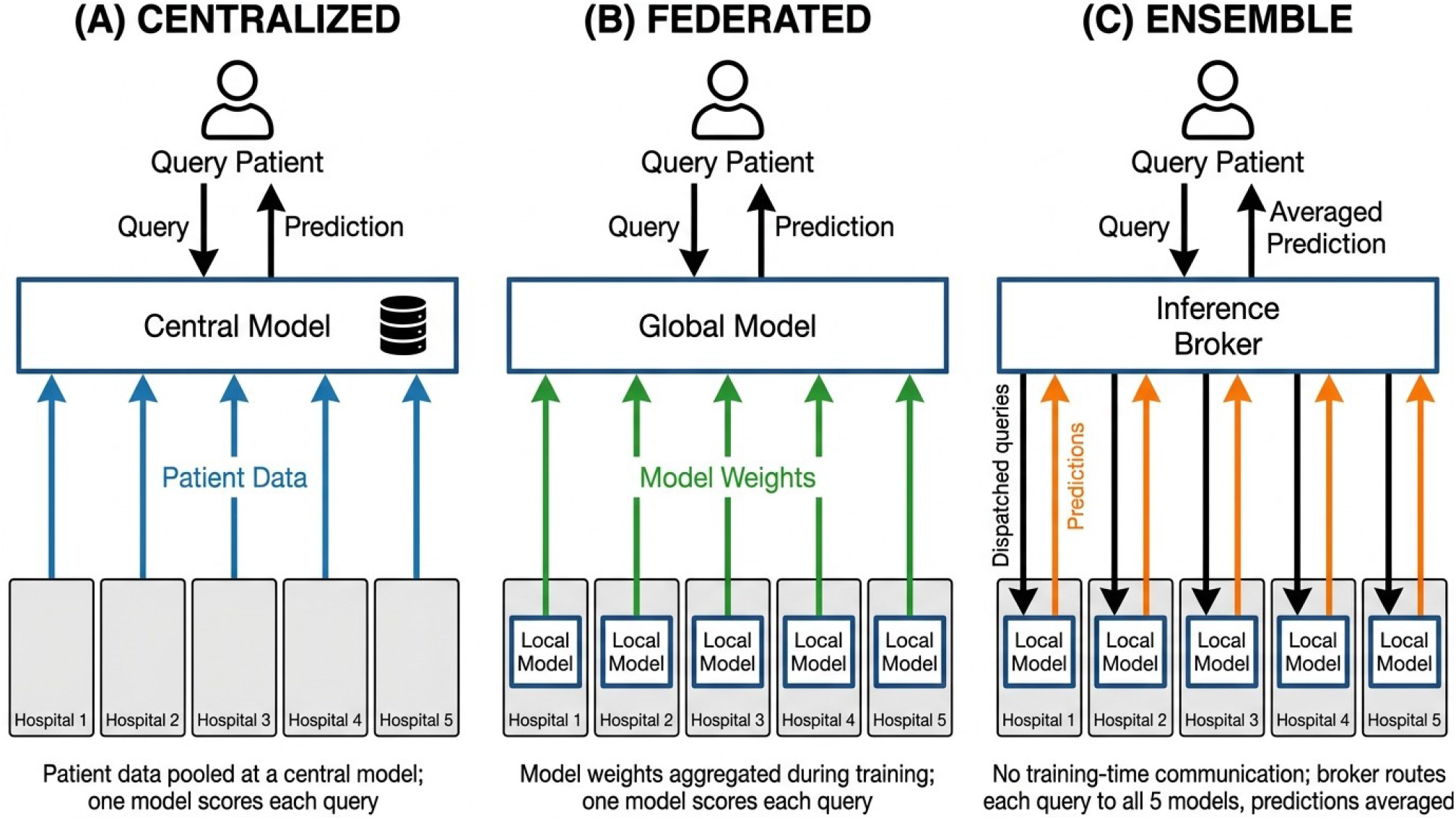
Three strategies for building multi-hospital EHR foundation models. All three strategies share the same three-tier structure: a query patient at the top, a coordinating component in the middle, and participating hospitals at the bottom. The strategies differ in what flows between hospitals and the coordinator. (a) Centralized training pools patient data from all hospitals into a Central Model; subsequent queries are scored by this single model. (b) Federated learning keeps patient data at each hospital and sends only model weights to a central aggregator across training rounds, which produces a Global Model that scores each query. (c) Inference-time ensembling trains each hospital’s model independently with no training-time communication; at query time, an Inference Broker forwards the query to all participating hospital models, collects their soft prediction probabilities, and returns the averaged prediction.

How these two strategies compare for autoregressive EHR foundation models has not been systematically evaluated. Prior federated-learning work in clinical prediction has focused on task-specific models, including EXAM for COVID-19 outcome prediction across 20 hospitals (8), rare-cancer boundary detection spanning 71 institutions (9), and bidirectional encoder representations from transformers (BERT)–style encoder architectures for medical concept embedding (10); whether federated learning can train generative, decoder-only EHR foundation models has not been established. Nor has it been established whether individual participating hospitals actually benefit from joining a collaborative foundation model or whether they would be better served training alone on their own data. Both questions are central to whether collaborative EHR foundation models will ever be built in practice.

In this study, we evaluate both strategies on a GPT-style EHR foundation model trained on MIMIC-IV, partitioned into five simulated non-IID hospital sites. We compare federated learning and inference-time ensembling against a centralized-training upper bound and quantify the benefit to individual hospitals of participating in an ensemble versus training alone. Our results show that a multi-institutional EHR foundation model is technically achievable without pooling data, that inference-time ensembling in particular imposes the lightest governance burden of any collaborative strategy, and that the majority of individual hospitals receive a measurably better model by participating than they could build on their own.

## RESULTS

### Centralized training provides the performance ceiling

We used MIMIC-IV (version 2.2), a publicly available critical care database from Beth Israel Deaconess Medical Center containing de-identified records for 422,739 patients (11). From this cohort we selected the 100,163 patients with at least two hospital admissions (enabling next-admission prediction) and split them 70/15/15 at the patient level into training (70,114), validation (15,024), and test (15,025) sets.

We trained a GPT-style decoder-only transformer (31.6 million parameters) on tokenized patient sequences comprising diagnosis, procedure, medication, and laboratory tokens, following the architecture of recent institution-scale EHR foundation models at NYU and UCLA (1,2). The model was trained with a multi-label next-visit prediction objective; at evaluation, sigmoid probabilities at a held-out visit’s position provide zero-shot predictions for all diagnoses in the target visit (see Methods for full architecture and training details).

To establish a reference for multi-site strategies, we first trained the model centrally on all 70,114 training patients. Centralized training achieved per-condition AUROCs of 0.75 to 0.85 across the five acute conditions tested (**Figure 2a**), consistent with the discrimination ranges reported by institution-scale models at NYU (1) and UCLA (2). Acute respiratory failure and heart failure were the most predictable (AUROC 0.84 each); sepsis was the least predictable but still well above random (AUROC 0.75). Overall AUPRC across the 2,590-code diagnosis sub-vocabulary was 0.376 ± 0.002 across three seeds (**Table 1, Figure 2b**). This establishes the performance ceiling that the multi-site strategies must be compared against.

**Table 1.** Summary of methods evaluated and overall performance. Each method is classified by what must cross institutional boundaries (training data, model weights, or predictions only) and what data volume trains each component. AUPRC is micro-averaged across the 2,590-code diagnosis sub-vocabulary; mean ± SD is reported across three seeds or Dirichlet splits where multi-seed data is available (single-seed entries reported without a ± term). Marginal and repeat baselines are deterministic.

| Method | Training data per component | What crosses institutional boundaries | Overall AUPRC | New-onset AUPRC |
| --- | --- | --- | --- | --- |
| Centralized | 70K (pooled) | Patient data | 0.376 ± 0.002 | 0.173 ± 0.001 |
| **FedAvg** | **70K (federated)** | **Model weights (rounds)** | **0.321 ± 0.005** | **0.148 ± 0.002** |
| Ensemble (5 sites) | ~14K per site | Predictions at inference | 0.291 ± 0.006 | 0.132 ± 0.003 |
| Per-site mean | ~14K per site | None (independent) | 0.251 ± 0.007 | 0.115 ± 0.003 |
| FedPer + LoRA (head ens.) | 70K (federated) | Model weights (rounds) | 0.242 ± 0.003 | 0.077 ± 0.001 |
| Linear bag-of-DX | 70K (pooled) | Patient data | 0.224 ± 0.000 | 0.029 ± 0.000 |
| FedPer (head ens.) | 70K (federated) | Model weights (rounds) | 0.218 ± 0.003 | 0.071 ± 0.003 |
| Repeat history | N/A | None | 0.161 | 0.003 |
| FedProx | 70K (federated) | Model weights (rounds) | 0.081 ± 0.003 | 0.033 ± 0.001 |
| Marginal frequency | N/A | None | 0.081 | 0.031 |

**Figure 2.**
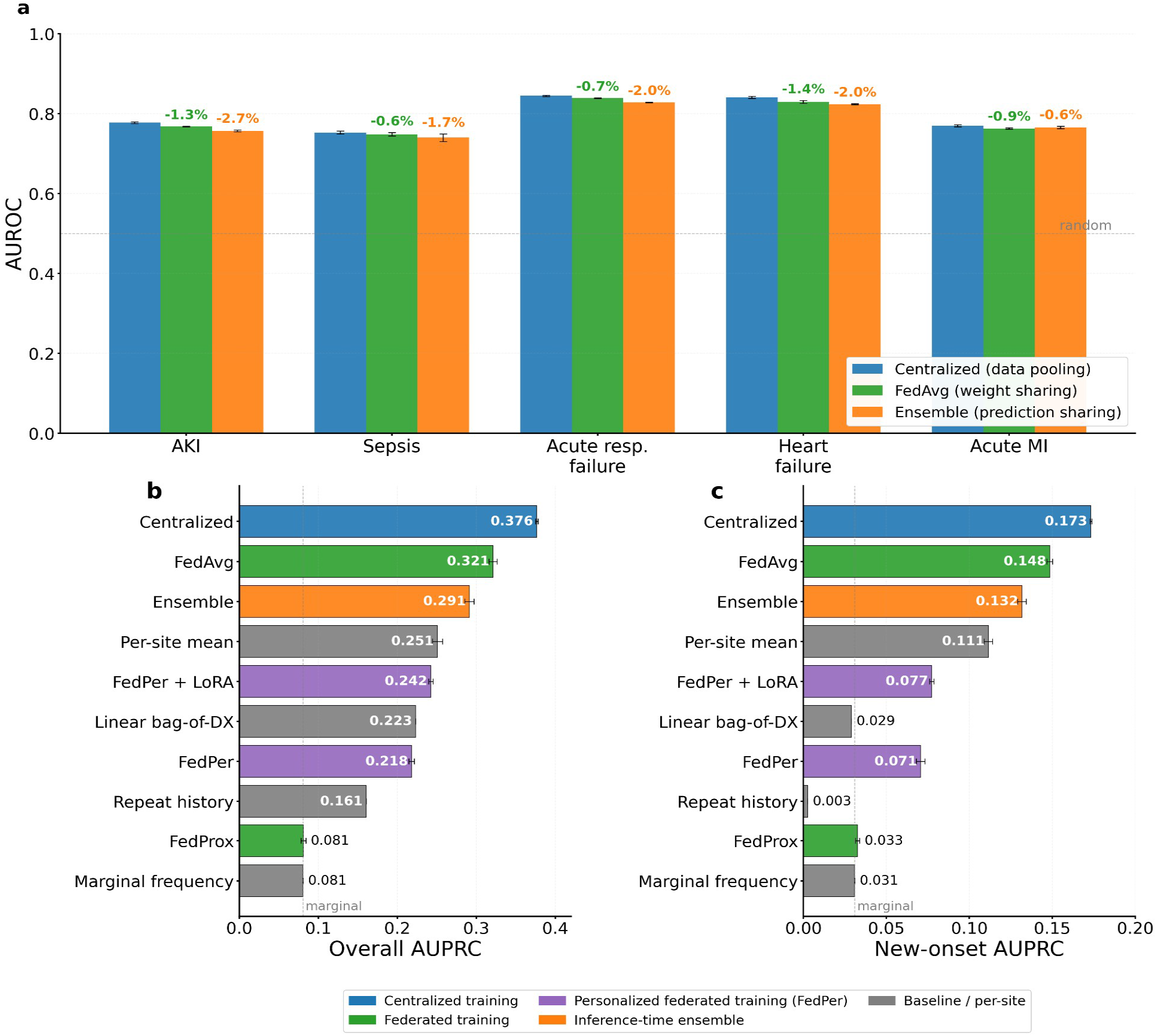
Multi-hospital strategies for EHR foundation models (simulated non-IID hospitals, Dirichlet alpha = 0.5). (a) Per-condition zero-shot AUROC for new-onset prediction across five acute conditions, comparing centralized training (blue), FedAvg (green), and inference-time ensembling (orange). Error bars show standard deviation across three seeds or splits. Percentage labels above the bars show the AUROC reduction relative to centralized training for the ensemble (orange) and for FedAvg (green). (b) Overall AUPRC across all methods tested. (c) New-onset AUPRC across all methods. Dashed line in (b) and (c) marks the marginal-frequency baseline.

### Federated averaging recovers most of centralized performance

We next applied federated averaging (FedAvg) to train the same model across five simulated non-IID sites (Dirichlet alpha = 0.5, 100 local steps per round, 20 rounds, three seeds). FedAvg achieved overall AUPRC 0.321 ± 0.005, recovering 85 percent of the centralized upper bound of 0.376 (**Figure 2b**). Per-condition AUROCs ranged from 0.75 to 0.84 (**Figure 2a**), recovering 98 to 100 percent of centralized performance. New-onset AUPRC was 0.148 ± 0.002 (**Figure 2c**), likewise 85 percent of centralized. The model converged smoothly across all three seeds, with validation loss matching the centralized trajectory within 6 percent.

FedAvg is not universally well-behaved across federated variants, however. FedProx, which adds a proximal regularization term to local training, produced AUPRC 0.081 ± 0.003 (**Table 1**), equivalent to the marginal baseline. This rules out FedProx for this class of generative pretraining objective, though the mechanism by which the proximal penalty collapses performance so severely is not fully resolved by our experiments. FedPer, which keeps the output projection per-site without averaging, achieved intermediate performance (AUPRC 0.218), below that of FedAvg; the site-specific output projections learned from only 14,000 patients each, whereas the averaged output projection in FedAvg effectively aggregated learning from the full 70,000-patient cohort.

### Inference-time ensembling is a viable alternative with a different governance profile

Federated averaging requires each participating hospital to share model parameter updates with a central aggregator. Many institutions may be unwilling or unable to share model weights, even with the patient data retained locally. Inference-time ensembling addresses this by training each hospital’s model independently and routing each query through an inference broker that collects and averages soft prediction probabilities from all participating models.

The inference-time ensemble across five independently trained per-site models achieved AUPRC 0.291 ± 0.006 (**Table 1**) and per-condition AUROCs of 0.74 to 0.83, recovering 97 to 99 percent of centralized AUROC (**Figure 2a**). The ensemble performed substantially better than any individual site model’s average (per-site mean AUPRC 0.251) and better than FedPer (0.218) or FedPer with low-rank adaptation (FedPer + LoRA, 0.242).

FedAvg outperformed the ensemble by 0.030 AUPRC overall (0.321 vs 0.291). This gap reflects the benefit of shared backbone training: FedAvg’s transformer receives gradient signal aggregated from all 70,000 patients across rounds, producing better internal representations than any independently trained site backbone. On per-condition AUROC, however, the two approaches were similar, differing by 0.01 to 0.02 per condition.

### Most hospitals receive a better model by joining the ensemble than by training alone

A collaborative foundation model is only adopted if individual hospitals benefit from participating. We therefore compared each hospital’s solo model, trained on only its own patients, against the ensemble that includes it. Based on 15 hospital-model evaluations spanning three independent Dirichlet splits of the training data, we estimate that approximately 87 percent of participating hospitals received a better model by joining the ensemble than they would have obtained by training alone (both on overall AUPRC and on new-onset AUPRC). Solo performance depended strongly on the hospital’s patient share in the network (Pearson r = 0.88). Only two hospitals in the sweep had solo AUPRC at or above the ensemble: one with 28,808 patients (41 percent of the network in its split) reached AUPRC 0.310, and one with 25,793 patients (37 percent) reached AUPRC 0.288. Every other hospital, with patient counts between 6,850 and 17,670 (approximately 10 to 25 percent of the network), performed worse alone than with the ensemble. For realistic clinical AI collaborations, in which no single hospital dominates the network, this means essentially all participating hospitals should expect a measurable improvement from joining.

### Ensemble performance is robust to site heterogeneity

To verify that ensemble results are not specific to one Dirichlet partition, we repeated the per-site training and ensemble evaluation across nine partitions (three splits at each of three concentration values, alpha in {0.5, 1.0, 2.0}). Ensemble AUPRC was stable across the heterogeneity range (mean 0.270 to 0.278 across alpha values, **Figure 3**), with the margin over per-site mean ranging from +0.025 to +0.038. The ensemble advantage was larger at lower alpha (more heterogeneous partitions), reflecting greater complementarity among specialized site models. Ensemble performance did not degrade as site heterogeneity increased, indicating that the strategy is appropriate for the realistic scenario in which different hospitals serve different patient populations.

**Figure 3.**
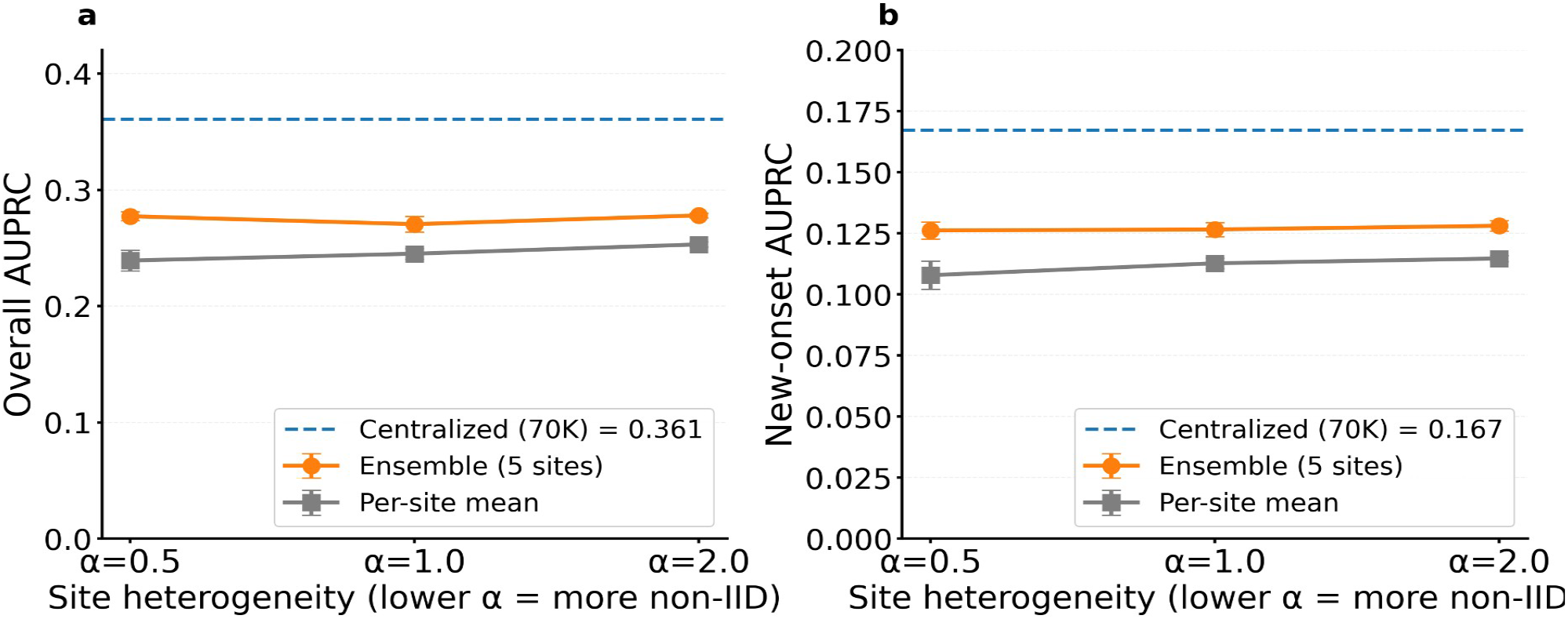
Robustness of ensemble and per-site performance to site heterogeneity. (a) Overall AUPRC and (b) new-onset AUPRC as a function of Dirichlet concentration alpha. Each point shows mean ± SD across three random splits. Dashed line marks the centralized reference. Ensemble performance is stable across the heterogeneity range, while per-site mean performance improves modestly with lower heterogeneity.

### All three strategies produce well-calibrated predictions

Discrimination metrics (AUROC, AUPRC) indicate whether a model can rank patients correctly, but clinical threshold-based decisions additionally require calibrated probabilities, meaning that a patient assigned a 10 percent risk should in fact develop the condition about 10 percent of the time. We therefore evaluated calibration (Brier score and expected calibration error, ECE) on the same five acute conditions, across three independent seeds or splits for each strategy (**Figure 4**). Brier scores averaged across conditions were 0.045 ± 0.019 for centralized training, 0.046 ± 0.019 for federated learning (FedAvg), and 0.046 ± 0.019 for the ensemble; ECEs were 0.009, 0.010, and 0.011 respectively. All three strategies achieved ECE values below 0.03 on every condition, and the reliability diagrams for each condition tracked the perfect-calibration diagonal closely across the range of predicted probabilities (**Figure 4**). Federated and ensemble predictions were therefore not only reasonably discriminative but also safe to use for clinical threshold-based decisions, with calibration essentially indistinguishable from centralized training. This finding matters because a common failure mode of collaborative models is that prediction probabilities drift from the true event rates even when discrimination is preserved; we did not observe this in our experiments.

**Figure 4.**
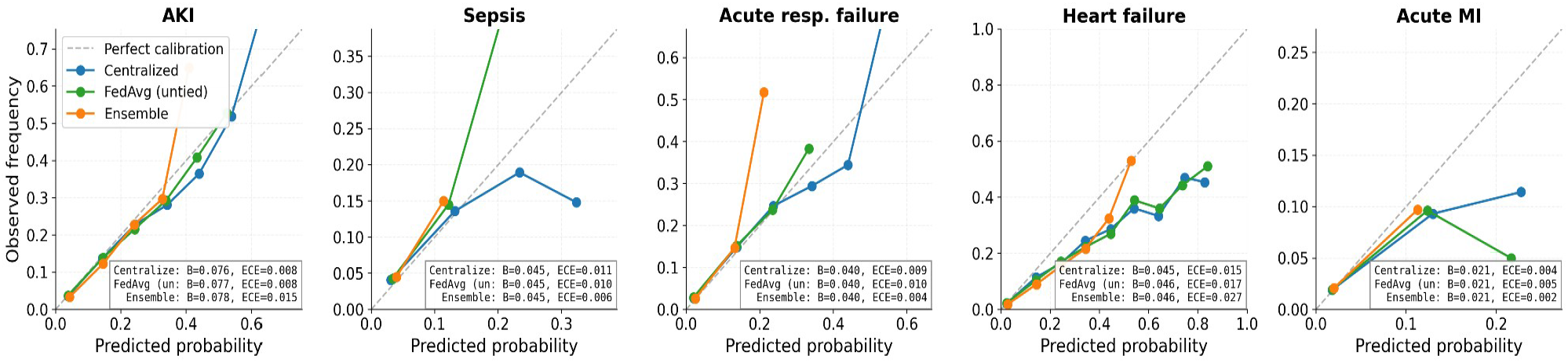
Calibration of centralized, federated, and ensemble predictions across five acute conditions. Reliability diagrams plot predicted probability (x-axis) against observed frequency (y-axis). Perfect calibration corresponds to points on the dashed diagonal. Each panel shows one condition with three curves for centralized training (blue), FedAvg (green), and the ensemble (orange), each averaged across three seeds or Dirichlet splits. Brier scores and expected calibration errors (ECE) are reported in each panel. All three strategies tracked the diagonal closely across the probability range and achieved ECE below 0.03 on every condition, indicating that federated learning and inference-time ensembling produce predictions that are not only discriminative but also well-calibrated for clinical threshold-based decisions.

## DISCUSSION

This study demonstrates that multi-institutional EHR foundation models are technically achievable without pooling patient data, and that individual hospitals stand to benefit from participating. No EHR foundation model trained across multiple institutions currently exists. If one is ever built, it will almost certainly be built under one of two regimes that avoid data transfer: federated learning or inference-time ensembling. We compared both on a simulated five-hospital partition of MIMIC-IV against a centralized upper bound and quantified their performance costs and governance implications.

The most practically important finding is that inference-time ensembling offers a viable collaborative path that requires no training-time coordination between hospitals and produces a measurably better model for almost every participating hospital than that hospital could build alone. Ensembling recovers 77 percent of centralized overall AUPRC (0.291 vs 0.376) and 97 to 99 percent of centralized per-condition AUROC across the five acute conditions we evaluated. Thirteen of fifteen individual hospital models (87 percent) were outperformed by the ensemble that included them; the exceptions were the two hospitals that received disproportionately large patient shares and could essentially match the ensemble independently. For any realistic multi-hospital collaboration where no single institution dominates the network, this means every participant can expect a better model by joining. The governance profile of the ensemble is narrow rather than globally private: no training data and no model weights ever leave any hospital, and no hospital sees any other hospital’s training data or model parameters at any point. At query time, however, the broker must be authorized to send the query patient’s tokenized history to each participating site’s model and collect back a vector of soft prediction probabilities, which it averages into the final prediction. This is a meaningfully different trust model from standard federated learning, but the scope of what leaves each hospital is limited to inference on the single query patient, rather than training-time parameter exchange across a whole cohort. Hospitals never share training data, never share model weights, and never need to coordinate training schedules, architectures, or hyperparameters. For research consortia, hospital networks, or commercial healthcare AI brokers working with partners who are unwilling or unable to share model weights, this is likely the only collaborative strategy that can be deployed at realistic scale.

Federated learning recovers more performance, 85 percent of centralized overall AUPRC (0.321) and 98 to 100 percent of per-condition AUROC, but requires hospitals to exchange model parameter updates at every training round. In contrast, FedProx — which adds a proximal regularization term to local training — collapses to the marginal baseline across the hyperparameter configurations we tested. For pretraining a generative EHR foundation model under federation we therefore recommend plain FedAvg, and explicitly discourage FedProx without further tuning.

When should each strategy be used? Centralized training should remain the first option where it is legally and institutionally feasible, for example within a single integrated delivery network, under a shared IRB, or inside a trusted research environment such as the National COVID Cohort Collaborative (N3C) (12). It achieves the highest performance and avoids the operational complexity of either alternative. Federated learning is appropriate for collaborations where all participating hospitals are willing to share model weights, such as established research consortia or single health systems with multiple hospitals; the 0.030 AUPRC advantage over ensembling reflects the benefit of shared backbone training across all 70,000 patients rather than per-site backbones trained on 14,000 each. Inference-time ensembling is appropriate for the broader class of collaborations where hospitals will not or cannot share model weights, such as broker-mediated partnerships spanning competitive health systems, international collaborations with differing regulatory frameworks, or institutions with restrictive model governance policies. In these settings, ensembling is likely to be the only practical path to a collaborative foundation model.

Several limitations bear on how these results should be interpreted. All experiments used simulated non-IID splits of a single dataset (MIMIC-IV); real multi-institutional heterogeneity includes differences in coding practices, EHR systems, patient demographics, and clinical workflows that Dirichlet allocation over ICD chapters does not fully capture. Many institutions map their EHR data to a shared common data model such as OMOP (adopted, for example, by N3C (12) and by the UCLA EHR foundation model (2)), which eases cross-site harmonization, but residual differences in unit conventions, value sets, local coding practices, and lab instrument calibration would still need to be handled in a real federated or multi-site ensemble deployment; we did not test any of this. Our model is relatively small at 28 to 31 million parameters, smaller than Columbia’s CEHR-XGPT (120 million) and NYU’s model (1.6 billion) (1,3); how these comparisons change at frontier scale is not settled. We evaluated five sites with approximately 14,000 patients each; behavior at very small per-site sizes, highly unequal sizes, or many more sites remains open. Our federated-learning comparison did not evaluate adaptive server optimizers such as FedAdam or variance-reduction methods such as stochastic controlled averaging for federated learning (SCAFFOLD). We did not evaluate subgroup fairness, which the FoMoH benchmark identifies as important for multi-site foundation models (5). The ensemble governance advantage we discuss depends on concrete deployment infrastructure (secure outbound scoring, query routing, or a trusted query aggregator); we describe the properties a suitable inference broker would need but do not ourselves provide one.

These limitations do not undermine the central practical conclusion. In a regulatory and institutional environment that has so far hampered multi-institutional EHR foundation model development, the experiments reported here show that such a model can be built without moving patient data across institutional boundaries, and that individual hospitals, of the size typical in real clinical collaborations, should expect to benefit from participating.

## CONCLUSION

Multi-institutional EHR foundation models can be trained without pooling patient data, and individual hospitals stand to benefit from participating. In simulated five-hospital collaborations on MIMIC-IV, inference-time ensembling recovered 97 to 99 percent of centralized per-condition AUROC while requiring no training-time exchange of any kind between participating hospitals; for 87 percent of individual hospitals, joining the ensemble produced a better model than the hospital could have trained alone. Federated learning recovered slightly more performance (98 to 100 percent of per-condition AUROC) but requires exchanging model parameter updates. Given that no published EHR foundation model has yet been trained across hospitals, and given the durable governance barriers to pooling patient data, these findings offer a concrete path forward: inference-time ensembling as the default collaborative strategy, with federated learning reserved for settings where model weights can be shared, and centralized training confined to the rare cases where data pooling is feasible.

## METHODS

### Data source and cohort

We used MIMIC-IV (version 2.2), a publicly available critical care database from Beth Israel Deaconess Medical Center containing de-identified records for 422,739 patients (11). We selected the cohort of 100,163 patients with at least two hospital admissions, enabling next-admission prediction. Patients were split 70/15/15 at the patient level into training (70,114), validation (15,024), and test (15,025) sets, stratified by admission-count bucket.

### Tokenization

Patient records were tokenized into sequences of clinical events organized by hospital admission. Each admission block contained diagnosis tokens (ICD-9 and ICD-10 codes collapsed to three-character prefixes, with version-specific prefixes DX9: and DX10:), procedure tokens (ICD procedure codes with PX prefix), medication tokens (first alphabetic token of drug name with MED: prefix, minimum training count 50), and laboratory tokens. Laboratory values were tokenized following the UCLA approach (2): for the top 200 most frequent lab item IDs, each value was assigned to a decile bin based on training-set empirical cumulative distribution functions. Only abnormal values (deciles 0, 1, 8, 9) were retained to focus on clinically meaningful deviations. The final vocabulary contained 6,256 tokens (4 special tokens, 2,590 diagnoses, 1,328 procedures, 809 medications, 1,525 laboratory tokens). Mean sequence length was 194 tokens per patient (median 129, maximum 1,024 after left-truncation of oldest admissions).

### Model architecture

We implemented a decoder-only transformer following Rajamohan et al. (1), with three key architectural features from that work. First, a two-level attention mask provided bidirectional attention within each admission combined with causal attention across admissions. Second, rotary positional embeddings (RoPE) indexed by inter-visit day delta rather than token position encoded continuous time intervals between admissions. Third, repeat-token decay regularization in the binary cross-entropy loss down-weighted chronic carry-forward conditions to prioritize new-onset events. The loss weight for each token was exp(-0.5 times r), where r is the number of previous admissions containing that token.

The model had 8 transformer layers, 8 attention heads, embedding dimension 512, feedforward dimension 2,048, and dropout 0.1, totaling approximately 31.6 million parameters. Laboratory tokens were included in the input sequence as contextual features but excluded from the prediction targets.

#### Input and output embeddings

The input embedding and output projection were implemented as separate (untied) linear layers. We tested both tied and untied configurations under centralized training and under FedAvg; untied performed slightly better on average across both settings (approximately 4 percent relative AUPRC centrally and 7 percent relative under FedAvg), so we report the untied configuration throughout the main text and **Table 1**.

### Centralized training

Training used AdamW optimization with learning rate 3e-4, weight decay 0.1, cosine annealing with 500-step linear warmup, gradient clipping at 1.0, batch size 32, and bf16 mixed precision on a single NVIDIA A40 GPU. Training ran for three epochs (6,573 optimizer steps, approximately 17 minutes wall time) across three random seeds (20260410, 20260411, 20260412).

### Simulated multi-hospital partition

To simulate realistic multi-institutional heterogeneity, we partitioned the 70,114 training patients into five sites using Dirichlet allocation over ICD chapter categories. For each ICD chapter (determined by the first character of each patient’s most frequent diagnosis code), we sampled a proportion vector from a Dirichlet distribution over five sites, then assigned patients of that chapter according to the sampled proportions. The headline experiments use Dirichlet concentration alpha = 0.5, which produces highly heterogeneous sites (one site enriched for cardiovascular patients while another is enriched for oncology patients), with site sizes ranging from approximately 10,000 to 18,000 patients. To assess robustness, we additionally swept alpha through {0.5, 1.0, 2.0} and resampled three random splits at each alpha (nine partitions total), retraining all per-site and ensemble models on each.

### Federated training

We evaluated federated averaging (FedAvg) and several personalization and regularization variants. Each round, every site initialized from the global model, trained for 100 local steps, and sent updated weights to the server for weighted averaging. We ran 20 rounds with server-side learning rate decay of 0.95 per round. All federated configurations used three random seeds at alpha = 0.5. The following variants were evaluated:

#### FedAvg

Standard federated averaging with untied embeddings (see above).

#### FedProx

FedAvg with an additional proximal regularization term (mu/2 times the squared distance from the global model weights) penalizing local parameter drift (7). Evaluated at mu = 0.1 across three seeds; additional tied-embedding FedProx runs at varying hyperparameter configurations gave consistent near-marginal results.

#### FedPer

Transformer blocks and input embedding averaged across sites; output projection kept per-site (not averaged, exempt from the proximal penalty).

#### FedPer + LoRA (two-phase)

Phase 1: FedPer for 20 rounds to build a shared backbone. Phase 2: frozen backbone with rank-8 LoRA adapters trained for 20 additional rounds.

### Inference-time ensembling

Each of the five sites independently trained a model on its own data for three epochs, with no inter-site communication during training. At evaluation, an inference broker forwarded each query patient to all five site models and their sigmoid output probabilities were averaged with equal weights to produce the final ensemble prediction. No training data, model parameters, or gradients were exchanged between sites at any point. We ran this protocol across three seeds.

## Evaluation

We evaluated all models on the held-out test set (15,012 patients with at least two admissions and diagnosis tokens in the target visit). The primary evaluation metric was per-condition AUROC for five acute conditions relevant to critical care populations: acute kidney injury (ICD-10 N17, ICD-9 584), sepsis (ICD-10 A40/A41, ICD-9 038), acute respiratory failure (ICD-10 J96), heart failure (ICD-10 I50, ICD-9 428), and acute myocardial infarction (ICD-10 I21, ICD-9 410). Per-condition evaluation included only at-risk patients (those without the condition in their prior history), with bootstrap 95 percent confidence intervals (1,000 samples). We additionally computed micro-averaged AUPRC across the 2,590 diagnosis sub-vocabulary as a secondary whole-vocabulary metric, separately for overall diagnoses and new-onset diagnoses. Error bars represent standard deviation across three random seeds or Dirichlet splits.

As baselines, we included marginal frequency (training-set population prevalence), repeat-history (diagnoses from the most recent prior admission), and a linear bag-of-diagnoses model trained with binary cross-entropy on patient-level diagnosis count vectors.

## Code and data availability

All code used to train and evaluate the models, generate the figures, and compile this manuscript is available at https://github.com/oelemento/federated-ehr-fm under the MIT license. The repository contains the full training and evaluation pipeline, SLURM submission scripts, the aggregate per-run evaluation metrics that drive every figure and table, and the manuscript source. MIMIC-IV (version 2.2) is freely available to credentialed PhysioNet users at https://physionet.org/content/mimiciv/2.2/; the repository does not redistribute MIMIC-IV data, derived patient-level artifacts, or trained model checkpoints, since these are derivative works of the credentialed dataset.

## Data Availability

MIMIC-IV v2.2 is freely available to credentialed PhysioNet users at https://physionet.org/content/mimiciv/2.2/. All code used to train and evaluate the models, generate the figures, and compile this manuscript is available at https://github.com/oelemento/federated-ehr-fm under the MIT license. The repository also contains the aggregate per-run evaluation metrics that drive every figure and table. No patient-level data, derived patient-level artifacts, or trained model checkpoints are redistributed.

## ACKNOWLEDGMENTS

O.E. is supported by UL1TR002384, OT2OD032720, and P01CA214274 grants from the National Institutes of Health, and by Leukemia and Lymphoma Society SCOR grants 180078-02, 7021-20, and 180078-01. Additional support was provided by the Englander Institute for Precision Medicine and the Department of Systems and Computational Biomedicine at Weill Cornell Medicine. Computation was performed on the Weill Cornell Medicine Scientific Computing Unit (SCU) and the Cornell Advanced Computing Center (CAC) Cayuga cluster. The author thanks the PhysioNet team for maintaining the MIMIC-IV dataset and the Beth Israel Deaconess Medical Center for contributing the source data.

## COMPETING INTERESTS

O.E. is co-founder and stockholder in Volastra Therapeutics, holds stock in Freenome, serves as a Scientific Advisory Board (SAB) member and holds stock options in Owkin, Harmonic Discovery, and Exai, and is an SAB member for Canary Biosciences. O.E. is receiving or has received funding from Eli Lilly, J&J/Janssen, Sanofi, AstraZeneca, and Volastra. None of these entities had any role in the design, execution, analysis, or reporting of this study.

## Notes

### Author Declarations

This study is a retrospective analysis of the publicly available, de-identified MIMIC-IV v2.2 dataset, distributed by PhysioNet. MIMIC-IV was released under the oversight of the Beth Israel Deaconess Medical Center (BIDMC) Institutional Review Board. Secondary use under the present analysis is governed by the PhysioNet credentialed-access Data Use Agreement. No additional institutional IRB review was required.

